# COVID-19-related testing, knowledge and behaviors among severe and chronic non-communicable disease patients in Neno District, Malawi: A prospective cohort study

**DOI:** 10.1101/2022.11.21.22282569

**Authors:** Haules Robbins Zaniku, Moses Banda Aron, Kaylin Vrkljan, Kartik Tyagi, Myness Kasanda Ndambo, Gladys Mtalimanja Banda, Revelation Nyirongo, Isaac Mphande, Bright Mailosi, George Talama, Fabien Munyaneza, Emilia Connolly, Luckson Dullie, Dale A. Barnhart, Todd Ruderman, the Partners In Health Cross-Site COVID-19 Cohort Research Network

## Abstract

**Objective:** To assess changes over time in COVID-19 knowledge, risks, symptoms, testing, and infection prevention practices among patients with complex non-communicable disease (NCD) receiving care at Neno District and Lisungwi Community Hospitals, Malawi.

**Design and participants:** We conducted a prospective open cohort study using telephone-based data collection among patients enrolled in NCD clinics. We conducted four rounds of data collection between November 2020 and October 2021.

**Setting:** Rural southwestern Malawi in Neno District which has a population of 150, 211 persons.

**Primary and secondary outcome measures:** We used descriptive statistics to characterize the population and assess COVID-19-related knowledge and behaviors. Linear and logistic regression models were used to assess significant changes over time.

**Results:** Across four rounds of data collection, the most commonly reported COVID-19-related risks among patients included visiting health facilities (range: 35-49%), attending mass gatherings (range: 33-36%), and travelling outside the district (range: 14-19%). Patients reporting having ever experienced COVID-like symptoms increased from 30% in December 2020 to 41% in October 2021, however, as of the end of study period, only 13% of patients had ever received a COVID-19 test. Overall, respondents answered about two thirds (range: 67-70%) of the COVID-19 knowledge questions correctly with no significant changes over time. Hand washing, wearing of face masks and maintaining safe distance were the most frequently reported strategies used to prevent spreading of COVID-19. Wearing of facemask significantly improved from 63% to 96% over time (p<0.001).

**Conclusions:** Households of advanced chronic disease patients reported accurate knowledge about COVID-19 and improved adherence to wearing of face masks over time. However, patients commonly visit locations where they could be exposed to COVID-19 and often experience COVID-like symptoms but are rarely tested for COVID-19. We urge the government and other stakeholders to increase COVID-19 testing accessibility to primary facility and community levels.

**Strengths and limitations of this study:**

1. Assessed COVID-19-related outcomes among a highly vulnerable group of patients in a rural African setting
2. Longitudinal follow-up allowed us to assess changes over time from December 2020 to September 2021
3. Data can inform COVID-19 infection preventive measures in a setting with persistently poor access to COVID-19 vaccines
4. The telephone survey was conducted among severe and chronic NCD patients in rural Malawi and is not generalizable to urban areas or rural populations without cellular service
5. Data was self-reported data and vulnerable to social desirability bias

## Introduction

People with pre-existing chronic diseases, including hypertension, diabetes mellitus, chronic renal failure, and ischemic heart disease, face elevated risk of severe illness, hospitalization and death from COVID-19 [1–3]. COVID-19 vaccines are effective at preventing severe illness [4,5] even among individuals with chronic diseases [1]. However, COVID-19 vaccination rates in low- and medium-income countries (LMICs) like Malawi remain low [6]. As of 11 March, 2022 Malawi had a total of 828, 080 people (4.4% of its population) been fully vaccinated against COVID-19 [7]. These low rates reflect a combination of inequitable distribution of global vaccines and vaccine hesitancy [8,9]. In this context of limited COVID-19 vaccination coverage, knowledge about COVID-19, access and acceptability of COVID-19 testing, and individual infection preventive measures continue to play a central role in mitigating the spread of COVID-19 [10,11]. These COVID-19 prevention measures, such as social distancing and mask wearing, are especially critical for unvaccinated chronic disease patients because of their elevated risk of severe disease with COVID-19 infection [10]. However, while extensive literature exists on COVID-related knowledge and practices in general population and among health care workers [12–14], fewer studies have focused on patients with non-communicable diseases (NCDs), especially in rural African areas [15].

In Malawi, surveys conducted in urban [16], primarily rural [17], and nationally representative populations [18] at the start of the COVID-19 pandemic (April-September 2020) found mixed evidence related to knowledge and practices surrounding COVID-19. For example, while many respondents understood that the virus could be spread through close contact with a COVID-19 case and through respiratory transmission [16,17] respondents also reported a number of misconceptions surrounding alternate routes of transmission and COVID-19 severity, especially in rural settings [17,18]. Similarly, although almost all respondents reported taking some steps to prevent COVID-19, increased handwashing was the most commonly adopted preventive measures, with markedly fewer people practicing social distancing or masking, especially in rural areas settings [17,18]. However, we known these patterns of COVID-19-related knowledge and prevention could be different among patients with non-communicable disease. These patients could have greater opportunities to learn about COVID-19 due to their frequent contact with the health system and be more highly motivated to adhere to COVID-19 prevention practices due to their elevated risk.

To better understand patients’ knowledge of COVID-19 prevention practices and vulnerability to COVID-19 infection, we conducted an open cohort study on COVID-19 knowledge, risks, symptoms and testing, and infection preventions practices among patients with complex NCD receiving care at clinics located at Neno District and Lisungwi Community Hospitals.

## Methods

### Study design and setting

We conducted a prospective open cohort telephone study among patients enrolled in NCD clinics at Neno District and Lisungwi Community Hospitals. These clinics provide the World Health Organization’s recommended Package of Essential Non-communicable Disease Interventions for first referral hospitals (PEN-plus). The hospitals are located in Neno District, which is in the southern part of Malawi with an estimated population of approximately 150,211 people [19] with 8,758 people that use electricity as their main source of lighting [20]. Partners In Health/Abwenzi Pa Za Umoyo (PIH/APZU), an international non-governmental organization, has worked in partnership with the Ministry of Health (MOH) to strengthen health systems and improve health outcomes in Neno district since 2007. In 2011, APZU/MOH established its first Chronic Care Clinic at Neno District Hospital, which provides longitudinal care for patients with an array of chronic diseases including HIV and common NCDs under one roof. In 2018, two PEN-Plus NCD clinics were established which provide outpatient services for severe and complex NCDs such as sickle cell disease, rheumatic heart disease and type 1 diabetes[21,22]. Currently, PIH/APZU and MOH supports care for over 4,000 NCD patients, including approximately 500 patients with severe and chronic NCDs.

### Study population

All patients who were a) enrolled in one of the PEN-Plus NCD clinics and b) had a telephone number included on their patient cards were eligible to participate. For patients who were under 18 years of age or critically ill, their adult caregiver gave consent to participate in the study and responded on their behalf. At the start of the study, out of 450 patients enrolled in advanced NCD as of December 2020, 105 had personal phones and an additional 50 had received phones as part of a PIH/APZU initiative to maintain continuity of care during COVID-19 pandemic, leading to an estimated telephone coverage of 34% for this study population (n=155). Due to the small size of this clinical population, we invited all patients who met our inclusion criteria in each round to participate.

### Data collection

We repeated the telephone surveys on a quarterly basis to track changes in patients’ COVID-19 related information over time, with the four rounds of data collection occurring in November-December 2020, March-April 2021; June-July2021, and September-October 2021. In each round, we updated the list of eligible patients with phone numbers and made up to five attempts on five separate days to contact eligible participants by telephone. Patients who were not able to be contacted after five attempts were referred to the clinical team to verify patient well-being via an in-person visit. The telephone interviews lasted approximately 25-40 minutes.

The data collection tools were adapted from a questionnaire developed by the Partners In Health Cross-Site COVID-19 Cohort Research Network, a team of clinicians and researchers from eight PIH-supported countries and methodologists from Harvard Medical School and Partners In Health-Boston. The questionnaire included modules on patient and respondent demographics and patient health history. We collected [17] data on patient’s COVID-19 related symptoms, COVID-19 risk factors and COVID-19 testing history, with caregivers responding on behalf of patients when necessary. We assessed the respondents’ COVID-related knowledge, which was assessed using questions adapted from Banda et.al [17] by asking respondents to report on their own COVID-19 related knowledge, whether they were the patient or a caregiver. We used an open-ended question to understand what action each household was taking to prevent COVID-19, and data collectors were asked to code responses according to a pre-defined list of select-multiple options. In the fourth round of the survey, questions of COVID-19 knowledge were replaced with questions on COVID-19 vaccination which will be reported in another paper. The questions were translated to “Chichewa” the local language and programmed in CommCare which is an open-source mobile application used to collect data. We recruited two enumerators to recruit study participants and administer each telephone survey interview using a CommCare application throughout the four cycles of data collection. The enumerators received a one-day training on how to administer the questionnaire and use CommCare before the first round of data collection.

### Data analysis

We described the population of respondents using frequencies and percentages for categorical variables and medians and interquartile ranges (IQR) for continuous variables. For each round, we reported the proportion of patients who experienced COVID-19-related risk factors, including visiting the health facility, attending a mass gathering, and traveled outside Neno, over the past two weeks. We reported the proportion of patients ever reporting covid-like symptoms which was defined as either loss of taste or smell or cough and at least one of fever, chills, muscle aches and shortness of breath. Similarly, we reported the proportion of patients ever experienced a COVID-19 test. We assessed respondents’ COVID-19-related knowledge by assessing whether they agreed or disagree with a series of six statements and reported the percentage of respondents in each round that answered a specific question correctly, with responses of “don’t know” classified as incorrect. We also calculated an overall summary of COVID-19 knowledge by dividing the number of questions answered correctly by the number of questions attempted by the respondent and assessed the proportion of respondents using the radio, Ministry of Health, friends and/or family, television, and social media as a source of knowledge on COVID-19. Finally, we reported the percentage of households engaging in various infection prevention control strategies. We visualized our data using bar charts and tested for significant difference between rounds using a regression model that included an indicator variable for each wave of the study and accounted for within-patient clustering over time. We used linear regression models for overall COVID-19 knowledge and logistic regression models for all other outcomes. Data were analyzed in Stata version 15.1.

### Patient and Public Involvement

Patients or the public were not involved in the design, or conduct, or reporting, or dissemination plans of our research.

## RESULTS

### Sociodemographic characteristics of patient and respondents

Over four rounds of data collection, we enrolled 192 participants, most of whom were enrolled in the first round (n=145, 75.5%) or third round (n= 42, 21.9%,). Sample sizes for each of the four rounds were 145, 131, 155, and 126, respectively. Of the 192 patients represented in the study, 101 (52.6%) were female and their median age was 50 years (IQR: 32, 68) with over one-third aged 60 years or older. Type 1 diabetes (n=50, 26.0%), hypertension (n=34, 17.7%), and chronic heart failure (n= 38, 19.8%) were the most commonly reported conditions. Most patients 114 (59.4%) had multiple conditions. Out of the 192 respondents, 126 (65.6%) were the patients themselves, the median age was 47 (IQR: 35, 62) and 111 (57.8%) were female (Table 1).

**Table 1.** Sociodemographic characteristics of patient and respondent (N=192)

|  | N (%) |
| --- | --- |
| Round of Enrollment |  |
| First Round (Dec 2020) | 145 (75.5%) |
| Second Round (Mar 2021) | 5 (2.6%) |
| Third Round (Jun 2021) | 42 (21.9%) |
| Fourth Round (Sept 2021) | 0 (0.0%) |
| Patient Sex |  |
| Female | 101 (52.6%) |
| Male | 91 (47.4%) |
| Patient Age |  |
| ≤5 years | 9 (4.7%) |
| 6-17 | 16 (8.3%) |
| 18-39 | 39 (20.3%) |
| 40-59 | 57 (29.7%) |
| ≥60 | 71 (37.0%) |
| Patient's Clinical Program |  |
| Type 1 Diabetes | 50 (26.0%) |
| Hypertension | 38 (19.8%) |
| Chronic Heart Failure | 34 (17.7%) |
| Type 2 Diabetes | 20 (10.4%) |
| Chronic liver disease/cirrhosis | 16 (8.3%) |
| Rheumatic/other heart disease | 17 (8.9%) |
| Chronic Kidney Disease | 10 (5.2%) |
| Other | 7 (3.6%) |
| Patient's number of conditions |  |
| 1 | 78 (40.6%) |
| 2 | 70 (36.5%) |
| ≥3 | 44 (22.9%) |
| Respondent type |  |
| Patient | 126 (65.6%) |
| Parent | 20 (10.4%) |
| Child | 23 (12.0%) |
| Spouse | 9 (4.7%) |
| Grandparent | 6 (3.1%) |
| Other | 8 (4.2%) |
| Respondent's Sex |  |
| Female | 111 (57.8%) |
| Male | 81 (42.2%) |
| Respondent's Age |  |
| 18-39 | 65 (33.9%) |
| 40-59 | 73 (38.0%) |
| ≥60 | 54 (28.1%) |

### COVID-19 risks, symptoms and testing among patients

In all four rounds of data collection, the most commonly reported COVID-19 related risks experienced by patients in the past two weeks were visiting the health facility (range: 35-49%), followed by attending mass gatherings (range: 33-36%) and travelling outside Neno (range: 14-19%). Visiting traditional healers, traveling outside Malawi, or being in contact with a COVID-19 case were rarely reported. Only the proportion of patients who reported visiting health facilities changed significantly over time (p<0.0183). In the first round, one-third (30%) of participants reported having ever experienced COVID-like symptoms and this increased to 41% in the fourth round (p<0.0132). Only 13% of the participants in the fourth round reported ever receiving a COVID-19 test at any point in the study and only one patient in the fourth round reported testing positive. Statistically, we found no significant change over time regarding receipt of COVID-19 testing (Figure 1).

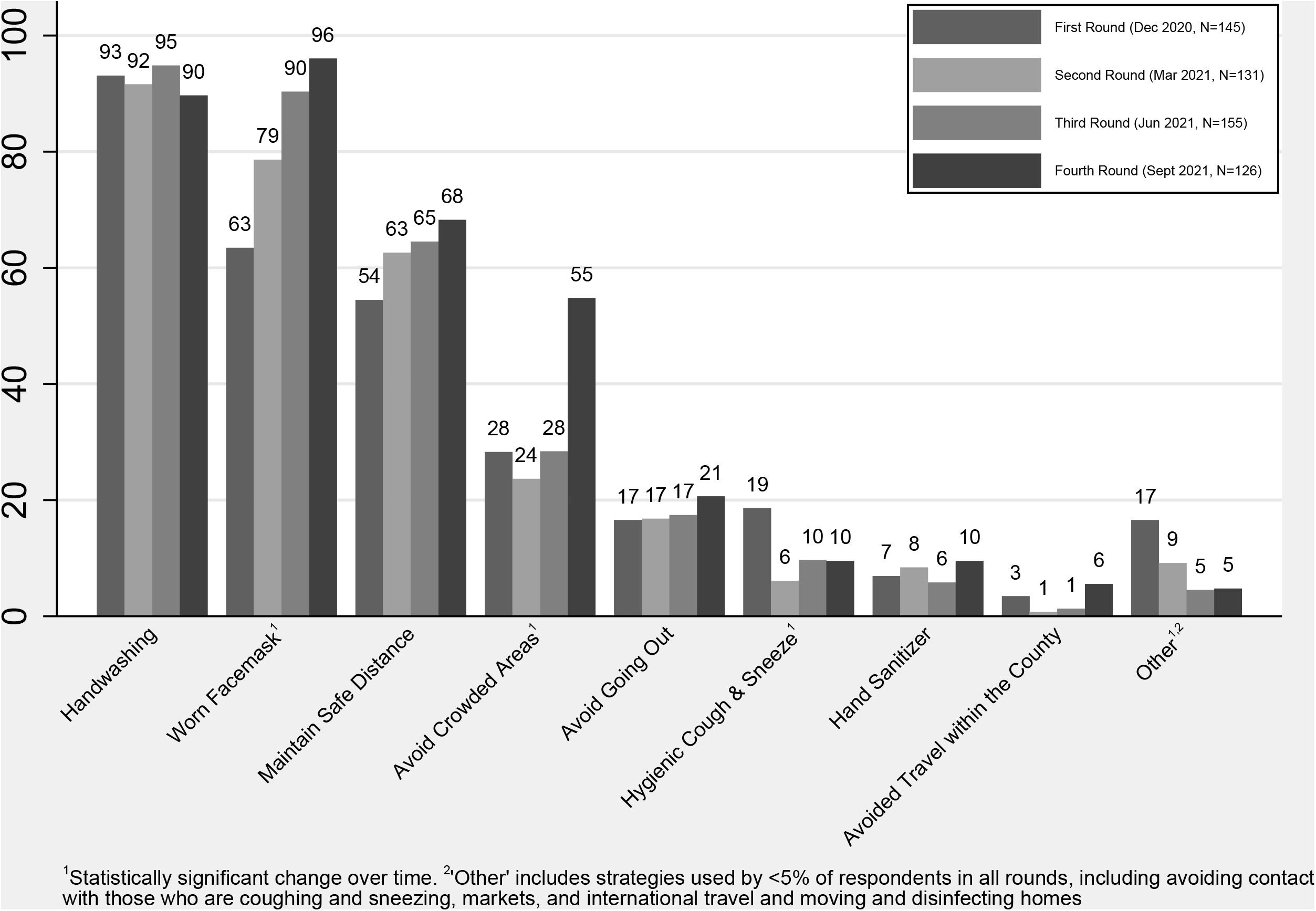

### COVID-19 Knowledge among respondents

Respondents answered about two third of the COVID-19 knowledge questions correctly with no significant changes over time (range: 67-70%). Overall, COVID-19 knowledge questions where “Agree” was the correct answer had a much higher percentage of respondents (range: 78-97%) answering correctly when compared to questions where “Disagree” was the correct answer (range: 22-31%) (Figure 2). Respondents most commonly cited the radio (range:81-85%), Malawi Ministry of Health (range: 79-86%), and friends or family (range: 43–59%) as sources of COVID-19 knowledge while television (range:7-8%) and social media (range:3-5%) were the least common sources of knowledge (Figure 3). The reduction in respondents citing family or friends as a source of COVID-19 knowledge reduced significantly over time (p<0.0093).

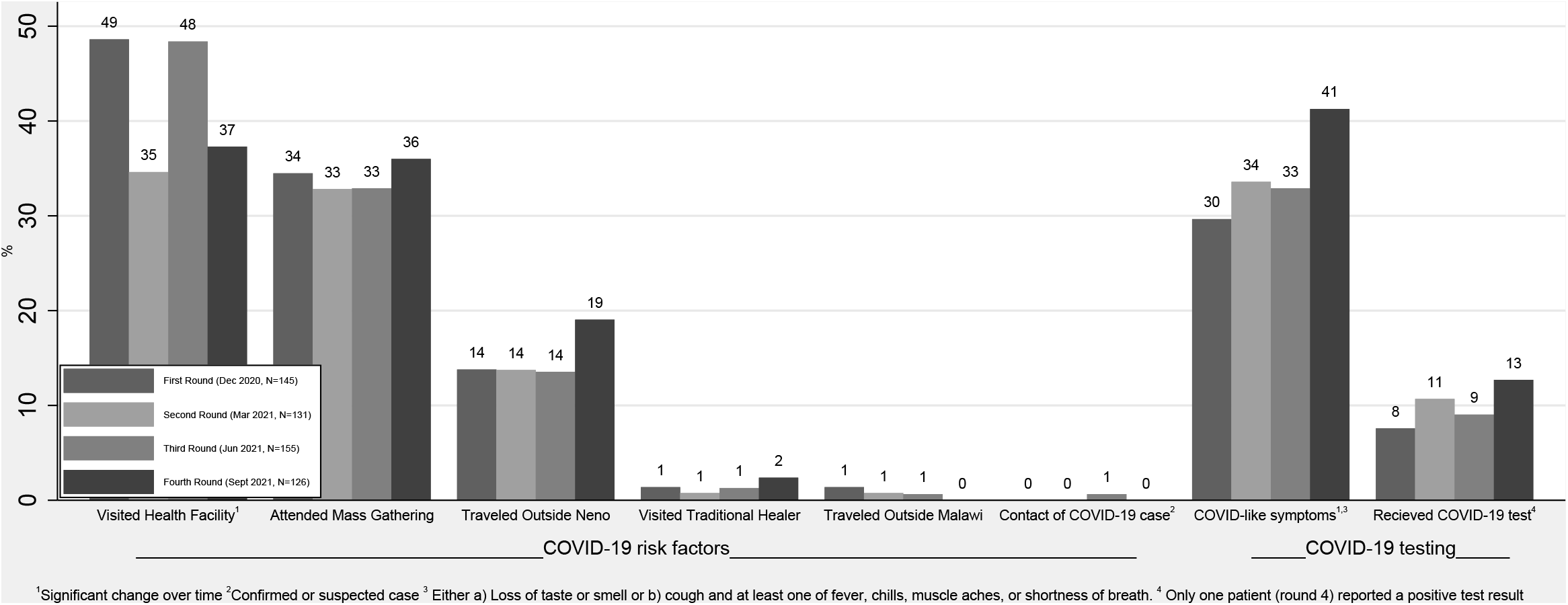

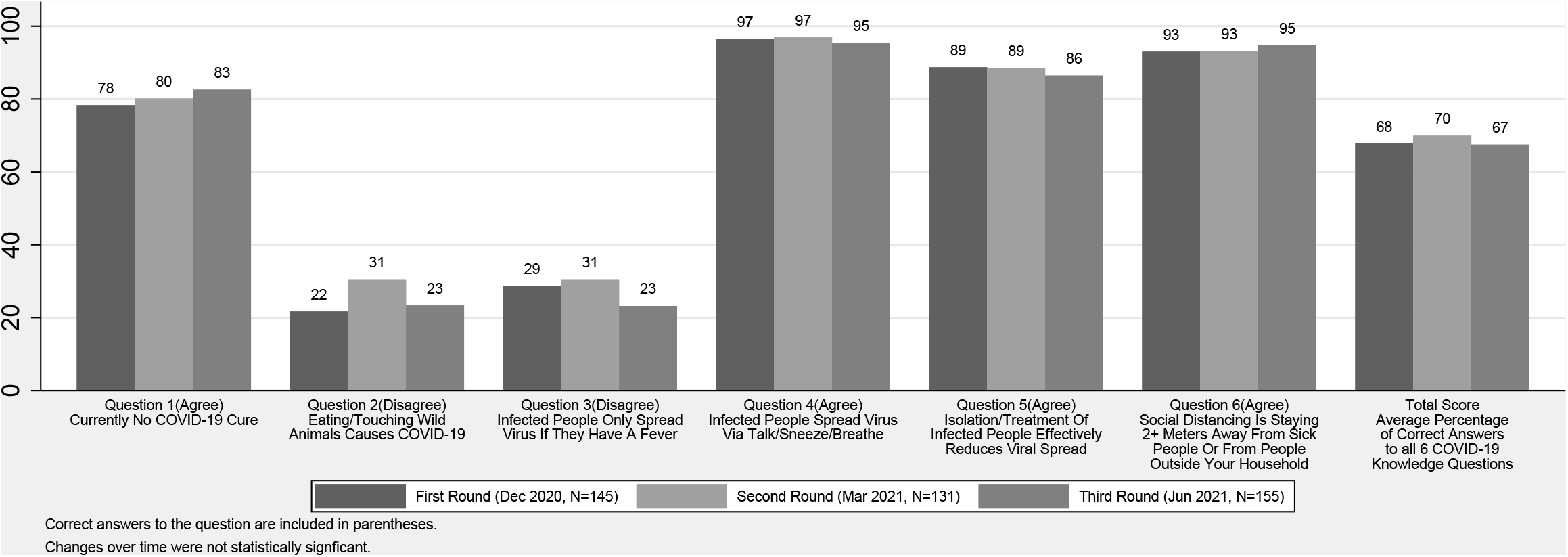

### Infection Prevention Strategy among households

The most commonly practiced infection prevention strategies were washing hands (range: 90-95%), wearing facemasks (range: 63-96%) and maintaining safe distance (range: 54-68%). The adoption of both wearing facemasks and avoiding crowded areas increased over time (p<0.001). Hygienic coughing and sneezing significantly decreased over time (p<0.0072). In all four rounds, less than a quarter of households reported avoiding going out, using hand sanitizer and avoiding within-country travel, with no significant changes over time (Figure 4). Use of infection prevention strategies classified as “other” dropped significantly from 17% to 5% over time (p<0.0013).

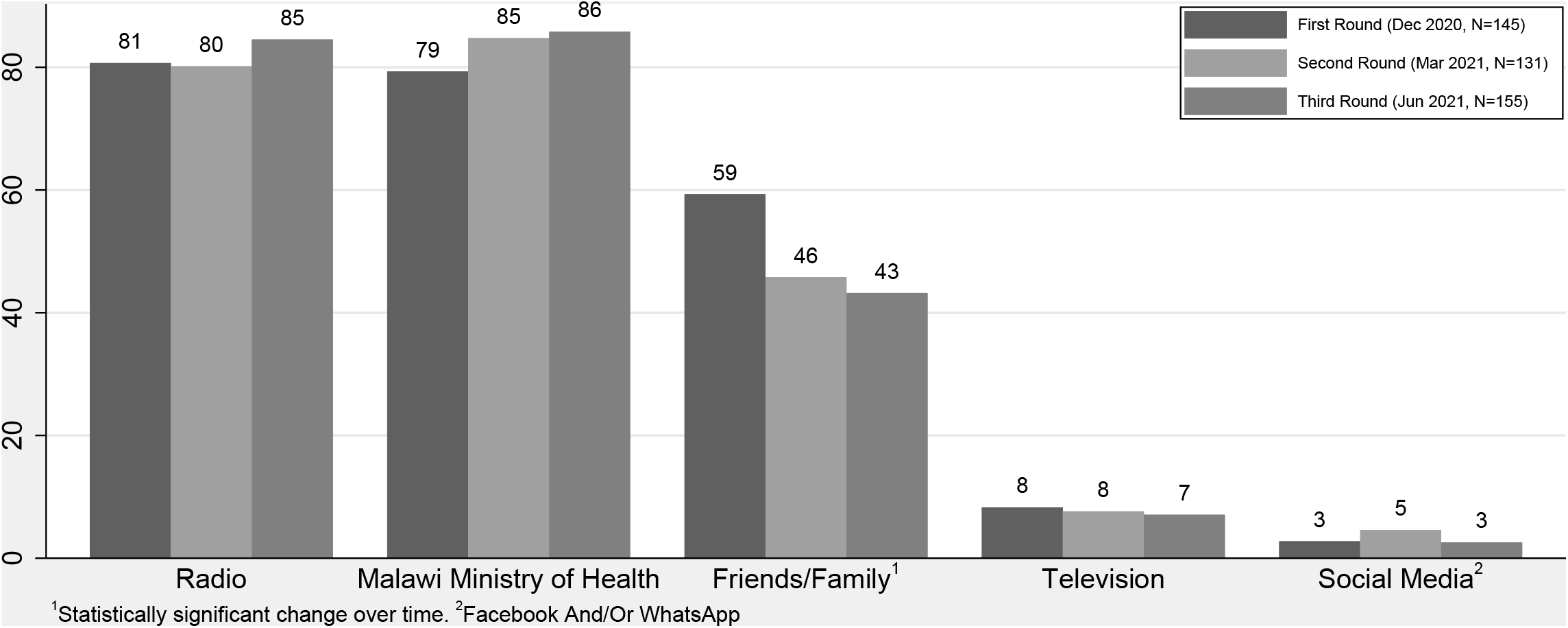

## DISCUSSION

In our prospective telephone-based open cohort study of COVID-19 patients with severe and chronic NCDs in rural Malawi, we found frequent exposure to COVID-19 risks with low COVID-19 testing. The participants had moderate knowledge about COVID-19, which did not improve over time; but high engagement in COVID-19 prevention activities, which did improve over time. Our study found visiting health facilities were the most common risk factor, likely reflecting high patient engagement in NCD care. However, visiting health facilities dropped sharply during rounds two (March-April 2021) and four (September-October2021). This reduction is likely due to reduced health service utilization during large waves of COVID-19, which occurred between early January and late March 2021 and between late June and late September 2021 [23]. Similar decreases in health services utilization during the waves have been reported in 20 countries [24]. Our findings suggest that our patients were skipping appointments in order to reduce their risk of COVID-19 at the expense of receiving appropriate care and treatment for their NCDs.

By the fourth round of our study, 41% or patients reported having ever experienced COVID-like symptoms but only 13% had ever received COVID-19 testing. Although this level of testing is sub-optimal, testing among our cohort in December 2020 (8%) was higher than what was reported by Banda et al. in a survey conducted among current and former residents of Karonga District, Malawi and their siblings in Nov 2020 (5.9%) [25]. Inadequate access to testing at health facilities is likely the major cause for this low coverage. Malawi relieved exclusively on PCR testing to diagnose COVID-19 from the start of the pandemic in March 2020 until rapid diagnostic tests (RDTs) received government approval in January 2021. Even after RDTs became available at Neno District Hospital, supplies were limited and RDTs were only used for symptomatic patients or oxygen saturation <92% [26]. Furthermore, since the RDTs were only available at the hospital and not at more local primary-level health facilities, patients with mild symptoms may have chosen not to get tested due to the distance to the facilities. Only one patient in our study reported testing positive for COVID-19. Given the low accessibility of testing and high COVID-related stigma [27], it is likely that this finding reflects widespread under-testing and under-reporting rather than low COVID-19 prevalence. Seroprevalence studies done in Malawi and other countries have reported increasing seroprevalence for SARS-CoV-2 over time (12.3%, 14.3%, 51.5%) [28–30]. Collectively, these findings underscore the importance of increasing the accessibility of decentralized COVID-19 testing, especially for vulnerable populations such as chronic disease patients and their household members.

Knowledge surrounding COVID-19 was moderate, but did not improve over time. Our findings were very similar to Banda et. al [25], who found that approximately 90% of rural respondents understood the virus was transmitted through respiratory droplets, approximately 70% knew that there was no effective treatment for COVID-19, but only approximately 75% knew that asymptomatic transmission was possible. However, in general, our patients had better knowledge than urban respondents from Li et al [16]. Because Li et al conducted household surveys while our research and Banda’s research used telephone-based data collection, these differences could reflect disparities in access to information among households that do not own a telephone. Since COVID-19 was recognized as a health risk in 2019, the Ministry of Health in Neno and its implementing partners have been disseminating information related to COVID-19 transmission, prevention and access to testing through a public address van, radio and health talks. However, the lack of improvements over time, especially surrounding persistent beliefs in misconceptions around COVID-19, suggest that additional efforts are needed to address and correct existing misinformation.

In general, household engagement in infection prevention strategies was extremely high. Increased handwashing was a particularly common response early in the pandemic, as has been reported previously in Malawi [16,25]. We also observed statistically significant improvements over time for household use of two key infection prevention measures: wearing face masks and avoiding crowded places Maintaining safe distances also increased overtime, although this difference was not statistically significant. Over the course of the COVID-19 pandemic, the government of Malawi has been implementing mandatory masks strategies when visiting health facility, markets and political gathering and previous research in Malawi has observed similar improvements in the use of facemasks and social distance over time [25]. However, as evidenced by the fact that a third of patients attended a festival or mass gathering in the past two weeks across all four rounds of data collection, adherence to these infection prevention strategies is not perfect.

Our study has several limitations. First, this telephone survey was conducted among severe and chronic NCD patients in rural Malawi and is not generalizable to urban areas, other rural populations, or individuals without access to cellular service. However, because the clinical team distributed phones to low-income NCD patients to support continuity of care during COVID-19, our study population may better capture low-income patients generally without mobile phones than other comparable telephone surveys. Second, all data was self-reported data, which can be prone to misreporting. In particular, reporting COVID-19 compatible symptoms or testing could be prone to recall error and may not be very specific or sensitive for diagnosis of COVID-19 cases. However, we do believe that our definition of COVID-19 compatible symptoms is sufficient to identify individuals who could benefit from additional COVID-19 testing and is a useful measure for understanding the current gaps in testing coverage. Similarly, social desirability bias could have impacted our study, especially since hospital staff could be perceived as authority figures by patients. This bias may have impacted reporting for COVID knowledge questions if patients were afraid to disagree with false statements. However, we tried to minimize this by recruiting enumerators who were not medical providers. Lastly our sample size was small, leading to imprecision in our estimates.

Despite these limitations, our study provides valuable information on COVID-19 related testing, knowledge and behaviors among highly vulnerable population patients with severe chronic NCDs in rural Malawi. Until COVID-19 vaccines are globally available in high-and low-income countries, these patients will remain vulnerable to COVID-19 infection and severe illness. Our research suggests a number of concrete steps that are still needed to support these patients. First, there is a persistent need for decentralization and increased capacity for testing in rural Malawi. Second, we urge the government, in conjunction with other stakeholders to consider providing social support for patients to follow COVID prevention measures. These should include, at a minimum, high-quality masks that can be used by patients with NCDs and their caregivers when they travel to health facilities or other crowded areas. Finally, there is a continued need for advocacy for increased availability of COVID-19 vaccines as well as educational interventions to encourage vaccine uptake among NCD patients and their household members.

## Data Availability

All data produced in the present study are available upon reasonable request to the authors

## Declarations

### Authorship contribution

DAB, TR, HRZ, and EC conceptualized the study. HRZ, TR, MN, IM, FM, and DAB contributed in the study design and protocol development. RN supported digitalizing the survey questionnaire. RN and MBA supervised data collectors and coordinated data collection. KV and KT managed data and conducted data analysis. HRZ, MBA and KV drafted the manuscript under mentorship from DAB. All authors critically reviewed and approved the final manuscript.

### Data sharing statement

No additional data available

## Acknowledgements

We would like to thank MoH and PIH/APZU leadership for the support during the implementation this study. We also would like to thank all patients in advanced NCD clinic and their guardians who took part in this study. This paper was developed under the Harvard Medical School Global Health Research Core and Partners In Health Writing Workshop, developed and facilitated by Dale A. Barnhart, Isabel Fulcher, and Bethany Hedt-Gauthier, with support from Donald Fejfar. Dale A. Barnhart and Kaylin Vrkljan provided direct mentorship to this paper as part of this workshop. We would also like to acknowledge the Cross-Site COVID-19 Cohort Technical Working Group, which is composed of the following members - Partners In Health/Boston: Jean Claude Mugunga, Donald Fejfar, Stefanie Joseph; Partners In Health/Haiti: Wesler Lambert, Mary Clisbee, Fernet Leandre; Partners In Health/Liberia: Prince F. Varney; Partners In Health/Lesotho: Melino Ndayizigiye, Patrick Nkundanyirazo, Afom Andom; Partners In Health/Malawi: Emilia Connolly, Chiyembekezo Kachimanga, Fabien Munyaneza; Partners In Health/Mexico: Zeus Aranda Remon; Partners In Health/Peru: Jesus Peinado, Marco Tovar; Partners In Health/Rwanda: Vincent Cubaka, Nadine Karema; Partners In Health/Sierra Leone: Foday Boima, Gregory Jerome; Harvard Medical School: Bethany Hedt-Gauthier, Isabel Fulcher, Dale Barnhart, Megan Murray.

## Ethics statement

We obtained an ethical approval from Neno District Health Research Committee (NDHRC/2020/015) and Malawi National Health Sciences for Research Committee (NHSRC/20/08/2594). Before the interview, we obtained oral consent utilizing a standard consent script from the respondent. All participants were at liberty to withdrawal from the study at any point in time during the interview process and were re-consented prior to each round. We de-identified the data before analysis to ensure anonymity. Data was stored on secure, password-protected servers and encrypted computers accessed only by trained study personnel.

